# Unlocking EMR Data to Track Physical Function Across the Continuum of Care

**DOI:** 10.1101/2025.06.22.25330086

**Authors:** Robin L. Marcus, Kelly Daley, Margaret A. French, Anne Thackeray, Erik H. Hoyer, Donna Beck, Daniel L. Young

## Abstract

**Introduction:** Physical function (PF) is a critical contributor to quality of life and healthcare value, especially for older adults at risk for functional decline following hospitalization. Tracking PF is essential for monitoring recovery, preventing adverse events, and improving care transitions. Despite the potential of electronic health records (EHRs) to enable tracking of PF, it is rarely tracked systematically. We present a case study on the extraction of PF data from EHRs for patients transitioning from hospital to homecare in a large health system, highlighting challenges and offering recommendations.

**Methods:** An expert consensus group identified data elements important to the measurement of PF. We then assessed the feasibility of extracting those elements from a single healthcare system. Working with Johns Hopkins Health System (JHHS) informatics and homecare leaders, we determined which elements were captured in the EHR and which were not feasible to extract within our resource constraints. We then requested a refined data list for adult patients during the project period. After validation, data were securely transferred to University of Utah Health (UUH).

**Results:** Data from 21,702 patients were included. Of 27 desired elements, 17 were successfully extracted. Elements were marked ‘present’ if documented at least once during admission, or ‘missing’ if absent. Administrative data had low missingness, while missingness for assessments of cognition and mobility performance in hospital were over 65% and assessments of PF capacity in home health were missing in over 80% of patients. However, 81.7% of those receiving home health rehabilitation had the expected mobility measure. Overall, 73% of patients had at least 75% of the extracted data elements.

**Conclusions:** To track PF effectively, begin with clear definitions, a targeted cohort, and relevant data elements. Collaboration with EHR, clinical, and billing experts is essential, as is upfront assessment of data availability and alignment with project resources.

## 1 INTRODUCTION

Physical function is an important independent contributor to quality of life and the ability to independently move about in one’s environment is an almost universal desire^1^. Mobility limitations are increasingly prevalent in older adults, affecting nearly 35% of those aged 70, and the majority of those over 85 years of age^2–4^. Older adults, and specifically those with recent hospitalization are at risk for declines in physical function that are often associated with poor health outcomes including re-hospitalization, diminished quality of life, loss of independence and death^5–8^. Among Medicare beneficiaries, older adults with mobility limitations have nearly $3000 higher total healthcare expenditures, $300 additional out-of-pocket expenses annually, and approximately 14 additional hospitalizations per 100 beneficiaries^9^ than those without such limitations. To better understand and address this important problem physical function must be tracked in a standardized way and across the continuum of care.

About 17% of U.S. adults over age 65 have a hospital stay annually^10^, over a third are discharged at a functional level lower than admission, and few regain preadmission function within a year^11^. Furthermore, patients often seek healthcare services when, in addition to symptom burden, some aspect of their daily physical function is interrupted^12^. This underscores the need to prioritize physical function as a meaningful patient outcome when delivering effective and accountable care. Patient important outcomes are key components of the healthcare value equation, essentially defining the overall quality of care patients receive ^13^. While physical function is a critical factor in assessing healthcare value, it is not routinely nor systematically tracked across the healthcare continuum ^14,15^.

Understanding how physical function changes over time and across settings is crucial because it can help to detect early signs of decline, enable timely interventions to prevent further deterioration, enhance quality of life, and reduce the risk of disability, falls, and other health complications. Loss of physical function, irrespective of how it is measured is a robust predictor of hospital readmission in older adults ^16^. Furthermore, physical function is a key factor that is diagnosis agnostic. This means that regardless of a patient’s primary medical diagnosis (e.g., heart failure, pneumonia), their ability to move and care for themselves is a fundamental component of their recovery and overall health ^17–20^. Because of this, tracking physical function can enrich not only individual patient care but the work of the entire healthcare and research team, irrespective of specific role. Electronic health records (EHR), widely adopted in the early 2000’s and now used in over 95% of hospitals and medical practices ^21^ have created an opportunity to improve tracking of health outcomes like physical function across time and healthcare settings. However, this potential is largely unrealized because of a lack of common language and standardized assessments for physical function; different disciplines often use disparate measures, making longitudinal tracking difficult.

Despite the promise of the EHR playing a key role in transforming healthcare into a more effective, and accountable system, use of the EHR to collect and extract data that can be used to drive clinical decisions, meaningful quality improvement, and clinical research remains elusive, particularly as it pertains to the measurement of physical function, a complex construct that encompasses multiple dimensions like mobility, balance, and endurance, and is influenced by a wide array of factors including cognition, social support, and the environment.

The purpose of this paper is to identify challenges and recommendations for using EHR data specific to the measurement of physical function and how it changes over time and across care settings, for clinical care and research. To illustrate these challenges, in this paper we provide an example of the process, barriers, and facilitators for acquiring key indicators of physical function from the EHR of a single, large healthcare system. This system was chosen because they have a robust and systematic process to collect and track physical function within the hospital and into home health care^22–28^. Specifically, we will 1) describe the process used to identify data elements impacting or directly measuring physical function, 2) the process to extract those metrics, 3) descriptively report the metrics we obtained, and 4) discuss challenges and recommendations for obtaining and using those metrics.

## 2 PROCESS

### 2.1 Identification of Data Elements Reflecting Physical Function

Our initial steps for identifying relevant data for understanding changes in physical function have been published previously ^1^, but for context we briefly summarize those activities here. We gathered a diverse group of individuals including patients, physical and occupational therapists, nurses, case managers, physicians, healthcare administrators, health informaticists, and health services researchers. These stakeholders represented three different health care systems and settings including the acute hospital, home health and skilled nursing. These stakeholders participated in a 4-round modified Delphi process. Additionally, two focus groups of the remaining stakeholders offered further insights.

The first three rounds were conducted remotely and anonymously to create a list of potential data elements and rank how critical and feasibly obtainable they might be. In this first round, we identified 99 potential data elements across nine domains: physical function, participation, adverse events, behavioral/emotional health, social support, cognition, complexity of illness/disease burden, healthcare utilization, and demographics. In the fourth, in-person round, we finalized the critical and what we perceived as feasibly obtainable list of data elements resulting in 27 individual data elements. Of these, we anticipated that 20 could be routinely accessed through the EHR (Administrative Data), while the remaining seven, not necessarily routinely collected, required either patient or clinician effort (Patient/Clinician Input Data) (Table 1). Focus group participants confirmed the results from the modified Delphi process. Importantly, this process confirmed that a broad data set was necessary to understand physical function.

**Table 1.** Original data elements desired vs. present and missingness.

| Data Element | Setting | Present | Missing (n, %) |
| --- | --- | --- | --- |
| **Administrative Data** |  |  |  |
| Age | Acute | 21702 | 0, 0.0 |
| BMI | Acute | 21638 | 64, 0.3 |
| Race | Acute | 21643 | 59, 0.3 |
| Sex | Acute | 21702 | 0, 0.0 |
| Zip Code | Acute | 21678 | 24, 0.2 |
| Insurance | Acute | 21284 | 418, 1.9 |
| Injurious Fall | Acute | Not obtained due to resource limitation |  |
| Deep Vein Thrombosis | Acute | Not obtained due to resource limitation |  |
| Hospital Acquired Pressure Injury (III or IV) | Acute | Not obtained due to resource limitation |  |
| Hospital Acquired Pressure Injury (Any) | Acute | Not obtained due to resource limitation |  |
| Hospital Acquired Pneumonia | Acute | Not obtained due to resource limitation |  |
| All Cause 30 Day Readmission | Acute | 21197 | 505, 2.3 |
| Hospital Discharge Disposition | Acute | 21702 | 0, 0.0 |
| MS-DRG | Acute | 21702 | 0, 0.0 |
| Case Mix Index | Acute | Not obtained due to resource limitation |  |
| Surgery | Acute | Not obtained due to resource limitation |  |
| ICU Stay | Acute | Not obtained due to resource limitation |  |
| Hospital LOS | Acute | 21702 | 0, 0.0 |
| Medication count | Acute | Not obtained due to resource limitation |  |
| Physical therapy, occupational therapy, speech, language pathology utilization | Acute | 21702 | 0, 0 |
|  | HH | 21702 | 0, 0 |
| **Patient/Clinician Input Data** |  |  |  |
| Cognition | Acute | 7400 | 14302, 65.9 |
|  | HH | 0 | 21702, 100.0 |
| CAM-ICU | Acute | 6776 | 14926, 68.8 |
| AM-PAC Mobility | Acute | 21042 | 660, 3.0 |
|  | HH | 3739 | 17963, 82.8 |
| AM-PAC Activity | Acute | 20857 | 845, 3.9 |
|  | HH | 780 | 20922, 96.4 |
| JH-HLM | Acute | 6322 | 15380, 70.9 |
| PLOF | Acute | 5767 | 15935, 73.4 |
| Pain interfering with movement | Acute | Not obtained due to resource limitation |  |
|  | HH | Not obtained due to resource limitation |  |

Possible unique patient n = 21702, present=data present based on CPT code billed to discipline specific cost center (Acute) or visit type (Home Health - HH), missing=data missing in report

### 2.2 Data Acquisition

After identifying these data elements, we aimed to test the feasibility of extracting them from a single healthcare system across a single specific transition of care, acute hospital to home health care, between July 2016 and March 2021. While these data were largely present in the EHR of this system, identification of their location and then acquiring them through database extraction while adhering to patient and business privacy and confidentiality concerns required substantial effort and collaboration between researchers and informaticists. The project was approved by the University of Utah Institutional Review Board (IRB 00104345).

#### 2.21 Initial Planning

We met with the Director of Rehabilitation Informatics of Johns Hopkins Health (KD) and the Rehabilitation Supervisor of Johns Hopkins Home Care (DB) to compare the list of desired data elements with those already being collected by providers in the system. We determined that 10 data elements were either not already part of routine care documentation or could not be extracted from the EMR given our available resources (i.e., money and time) (Table 1). We also discussed the documentation practices and the general location of the remaining items in the EHR (e.g., flowsheet location). After this, the Director of Rehabilitation Informatics of Johns Hopkins Health met with representatives from the Johns Hopkins Hospital (JHH) Core for Clinical Research Data Acquisition (CCDA) to finalize the list of data elements and their respective locations. This allowed the JHH CCDA to provide an estimated budget for the work which was then reviewed and approved by the research team.

#### 2.22 Formal Data Acquisition

A data request was submitted to the JHH CCDA using a list of data field names and Epic Flowsheet IDs for either JHH acute or Johns Hopkins Medicine Home Care (JHM). We included adult patients discharged home with home health services from JHM during the project period. The Flowsheet IDs were gleaned by the Director of Rehabilitation Informatics at JHH from the front-end builder view of Epic. The CCDA data analyst used a multi-row SQL query to extract the requested data elements. That data was validated by technical review prior to being sent to the Director of Rehabilitation Informatics at JHH for clinical validation. Clinical validation consisted of front-end chart review of the identified data fields. Due to established research relationships with the acute care team, clinical validation of data from JHH acute care was more easily achieved. In contrast, since this process was new for the Home Care team, validating Home Care data required the recruitment of a home care supervisor (DB).

The data were comprised of 27 data elements from more than 35 discrete data fields. The discrete data fields came primarily from flowsheets and ‘smart data elements’ and included calculated assessment scores (e.g. SLUMS score), custom list values (e.g. bathroom set up), and text fields (e.g. type of home). Each data element was associated with a unique deidentified patient identifier, a unique deidentified encounter number, and the date/time of documentation.

The data was saved in a secure, HIPAA-compliant folder on a JHM virtual desktop. Once validated, fully extracted, and prepared, the data files were encrypted and transmitted via secure file transfer protocol to University of Utah Health (UUH). The password was shared with the UUH research team to unencrypt and open the files. UUH Information Technology allowed secure file transfer across their firewall from the Johns Hopkins server IP address via the Secure Socket Shell Protocol (SSH).

## 3 RESULTS

### 3.1 Description of the Acquired Data

#### 3.1.1 Overview of the Cohort

Data from 28,658 inpatient admissions for 21,702 individual patients were included in the initial data files. There were 4,332 individuals with more than one admission. For these individuals, we examined only the first admission during the study window, resulting in one inpatient admission for each of 21,702 individuals. We then identified the corresponding home health episode of care that followed that inpatient admission.

#### 3.1.2 Data Elements Omitted

We were unable to obtain 10 of the 27 data elements identified by the expert panel due to resource constraints (Table 1), although with unlimited resources, we likely would have been able to extract data related to these items. These included injurious falls, deep vein thrombosis, hospital acquired pressure injury, hospital acquired pneumonia, medications, case mix index, surgery type, and pain. For example, we found that *pain* was documented in many different ways in the medical record. Some of these included a Visual Analogue Scale (0-10), the FACES Wong-Baker scale (0-10 with corresponding diagrams of faces), the Behavioral Pain Scale which evaluates facial expressions, body movements, muscle tension and compliance with the ventilator, and lengthy subjective descriptions related to body area and aggravating or relieving factors. Reliably acquiring a measure of pain for each patient would have required additional resources like natural language processing and significantly more budget than we had anticipated when designing our original project. Another example of resource intensive data are *medications*, which we also knew were present, but would have required extraction logic of some complexity. Medications are prescribed through inpatient and outpatient orders but delivered via the “MAR” (medication administration record) for inpatients and filled by outside pharmacies for outpatients. Medication orders are often started, changed, stopped, and restarted, particularly in acute care. Inpatient lists may include many medications, dosages, routes, and frequencies during an admission. Medication reconciliation records merge home medications with new hospital medications but are sometimes incomplete or missing. We also found another data element, *injurious falls*, was not documented in the regular EHR but in a separate database for risk management and reporting and not available to us.

#### 3.1.3 Data Elements Extracted

Our goal was to assess the feasibility of obtaining the identified data elements related to physical function from the EHR, not to describe the functional status of these individuals. As a result, when examining the data we determined if each individual had at least one instance of each data element documented within the appropriate setting of care. If there was one instance of the data element, we labeled it as ‘present’ for that individual; if there was not a value documented for the given data element, we labeled it as ‘missing’. To examine the feasibility of our data extraction, we report the proportion of individuals with each data element ‘present’ or ‘missing.’ We were able to locate and had resources to extract 17 of the 27 individual data elements we wanted (10 administrative, 7 patient/clinician input in the inpatient and/or home health settings). The number present and number and percentage of missing records for each element is listed in Table 1.

For many administrative data elements, the missingness was below 1%; however, for assessments of cognition (Cognition, CAM-ICU) and mobility performance in the hospital (JH-HLM) missingness was over 65% and assessments of physical function capacity (AM-PAC Mobility, AM-PAC Activity) in home health were missing in over 80% of patients. This does not reflect a failure of health care providers to do their job but rather a policy and practice decision that these assessments are not needed for many patients. While we did not expect all patients to have all data elements, some of these numbers, (e.g., AM-PAC Mobility, AM-PAC Activity in the home health setting) were lower than expected. We then performed a secondary analysis with the subset of patients who received home health rehabilitation services. Only those who received home health rehabilitation services were expected to have an AM-PAC basic mobility measure and, in fact 81.7% (3553/4345) did.

Of note, when examining rehabilitation utilization (i.e., physical, occupational, and speech therapy), we assumed that if an individual had an evaluation CPT code for a specific rehabilitation discipline billed, they received care from that discipline; on the other hand, if there was no evaluation code billed, we assumed that the individual did not receive therapy from that discipline. In both cases, we had information about utilization. In other words, even if no CPT was billed it was informative, and we considered this data element to be ‘present.’

The involvement of rehabilitation therapists (physical and occupational therapists and speech language pathologists) is relevant to changes in physical function and so was among the data we sought to acquire. Using therapist procedural codes as indicators of rehabilitation therapist visits, we were able to quantify the involvement of each of these disciplines in each patients care. We summarize this involvement as ‘utilization’ in Table 2. This summary indicates what proportion of patients received at least one visit from each of these rehabilitation therapists during their admission or home health episode of care. Most patients in the acute hospital were seen by a physical therapist but less than half by an occupational therapist or speech language pathologist. In home health a large majority of patients were not seen by any rehabilitation therapist.

**Table 2.**
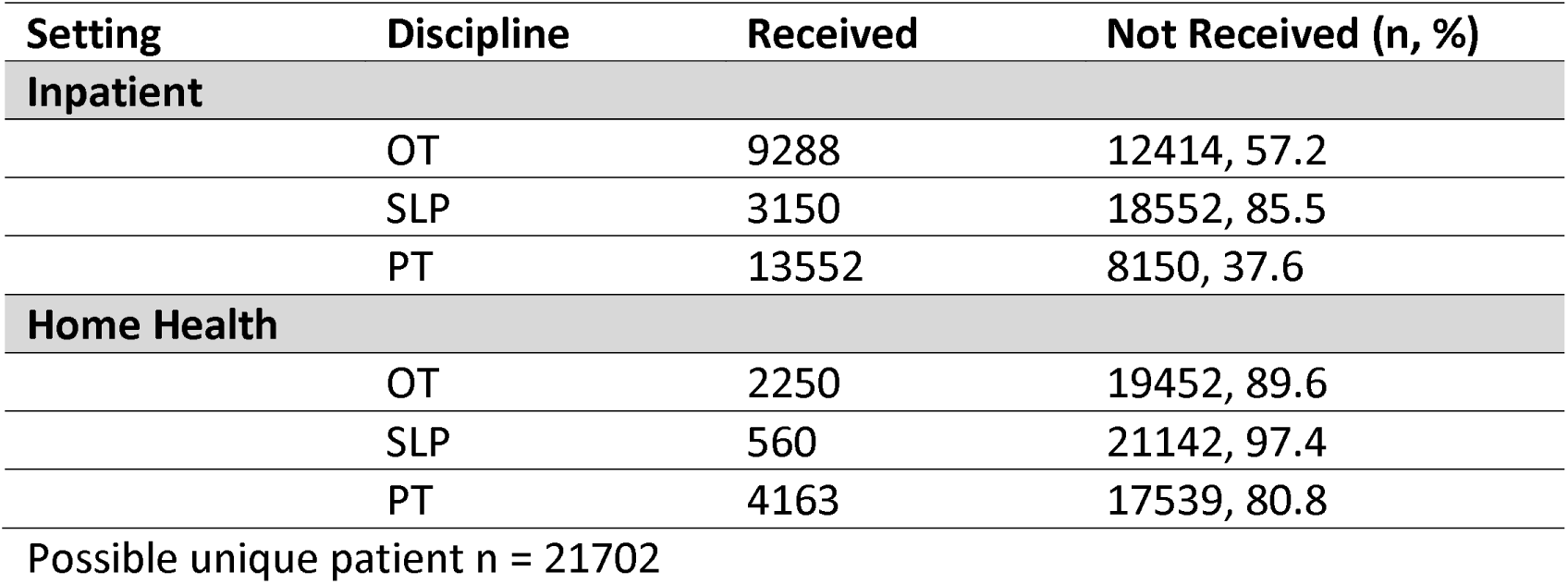
Utilization.

#### 3.1.4 Completeness of Extracted Data

The expert panel identified all these data elements as important for understanding physical function. As a result, we were interested in the proportion of data elements found for each individual. In other words, we examined how ‘complete’ the measures of physical function were in our cohort. The 17 data elements that we attempted to extract were represented by 20 unique data fields because certain data elements, such as cognition, were represented across multiple data fields in the EHR. Figure 1 shows the distribution of patients based on the proportion of these 20 fields successfully obtained. Seventy three percent (15846/21702) of the individuals in the cohort had at least 75% (15/20) of the data elements we extracted, while no one had all the data elements.

**Figure 1.**
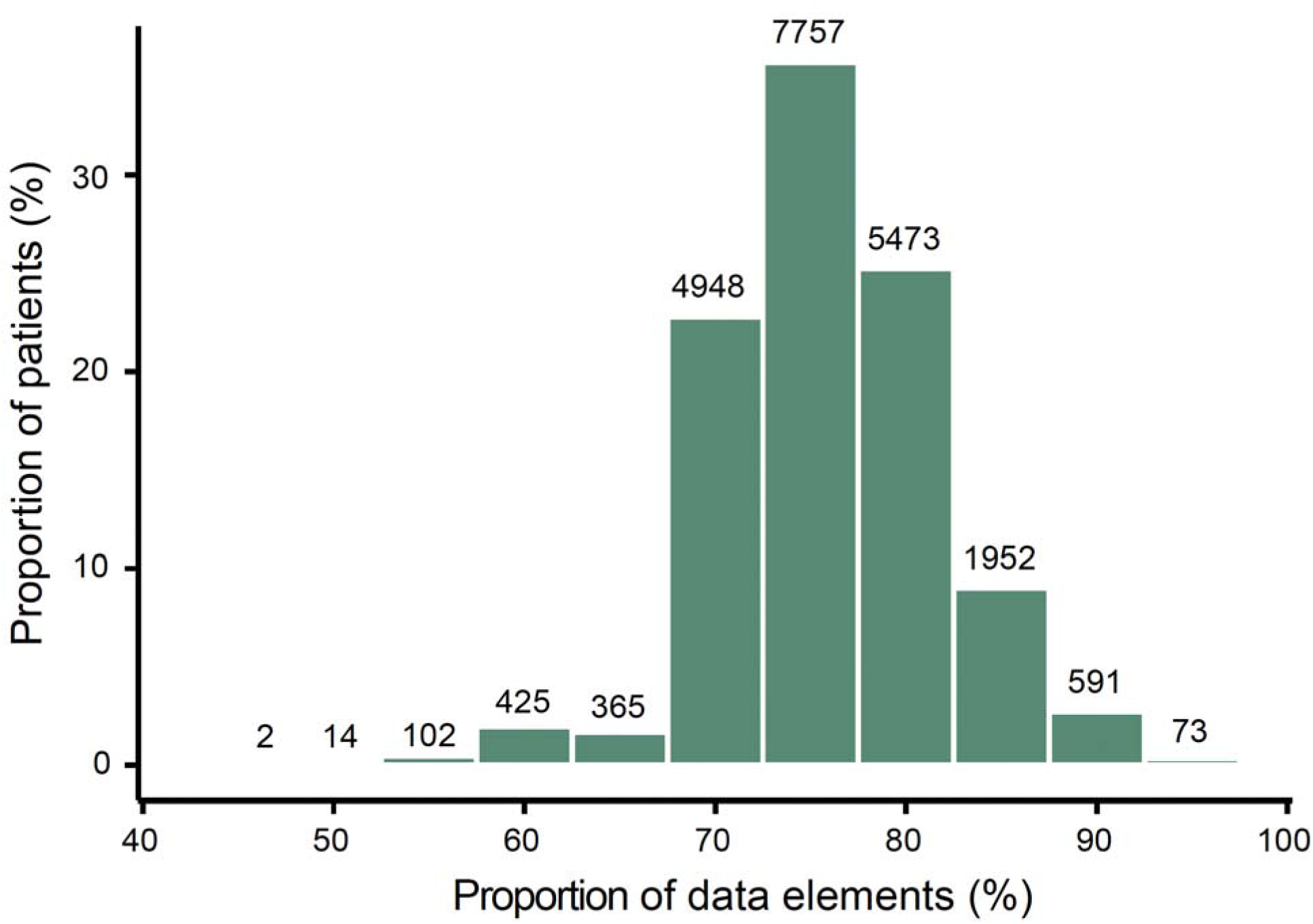
Proportion of data elements collected per patient

### 3.2 DISCUSSION

#### 3.2.1 Primary Challenges

After reaching consensus on a core set of data elements essential for measuring physical function—elements deemed both important and feasible to collect and extract from the electronic health record (EHR)—this project set out to identify the challenges and to make recommendations for obtaining these data for clinical care and research. Drawing on the EHR of a single large healthcare system, spanning the transition from acute care to home health care, we partially achieved our objective. The challenges we encountered fell into several key categories: data generation, storage, and extraction; data cleaning and validation; defining the patient cohort; difficulties linking data across care settings; and planning for adequate resources to manage the types of data involved. We learned that despite our planning and prior experience we were underprepared for some of these challenges. Below we discuss specific challenges we faced in the current project.

Even when data are entered into discrete fields, several factors make acquisition of usable data challenging. A major challenge was understanding how, when, and by whom data were generated, and where they were stored in the EHR. We observed that different patient care providers often documented the same clinical construct using different data elements and at varying time intervals, resulting in a complex dataset. Additionally, many critical data elements represent multidimensional constructs that are not collected in the same way by all providers such as cognition, pain, changes in medical status, or medication use. These are documented by different providers at different times and in different ways.

We encountered significant challenges in cleaning and validating data due to some of the issues discussed in the previous paragraph. This highlighted to our team the importance of building in time and effort for determining clearly defined operational definitions for our key constructs. For example, “primary diagnosis” appeared in the EHR in multiple forms. We ultimately chose to use the MS-DRG classification; however, this decision may have led us to document the primary diagnosis differently than if we had used an alternative source. We also faced challenges with identifying the timing of events that we suspect came from differences between date counts and day counts. Formatting discrepancies like these also require proactively allocating time and effort to resolve them. Additionally, we encountered challenges in extracting administrative data related to clinical events such as falls, DVTs, hospital-acquired pressure ulcers, and pneumonia. We suspect that many of these issues could have been avoided by spending more time and resources establishing clearer operational definitions and assessing data availability before initiating extraction. Identifying what data is needed, its source, and its accessibility—early in the project—would save considerable time and resources.

Common data models, such as PCORNet (Patient-Centered Outcomes Research Network), may ultimately facilitate the cleaning and validation process. These models standardize health data formats across participating healthcare systems harmonizing terminology and establishing uniform data definitions, formats, and coding systems across different healthcare organizations ^29^. The current challenge is that many of the variables identified by our stakeholders as essential data elements for physical function are not well represented in many common data models ^30^.

Cohort definition was another critical step, especially for data validation. Because not all patients receive the same services or interact with the same provider types, we needed to determine which patients were expected to have specific data elements. For example, only some home care patients receive rehabilitation services, and only rehabilitation providers collect our target mobility and activity measures in that setting. In contrast, hospital nurses, in addition to rehabilitation therapists, collect our target mobility data making it available on nearly all patients during hospitalization. Thus, tracking physical function into home health reduced our sample size more than expected. Moreover, a subset of patients likely received out of network homecare services, reducing the sample size even further. Additionally, clear indicators of admission and discharge from both hospital and home care were not consistently defined or recorded. While our project utilized data from a single healthcare system, we underestimated the complexity involved in connecting patient data between acute hospitalization and home health care.

The barriers we experienced to obtaining all the data we were initially interested in stemmed primarily from budget limitations. In particular, we underestimated the effort required for informatics resources. These challenges highlight the importance of collaboration with individuals who have deep knowledge of clinical documentation, EHR access, and billing practices—namely, clinicians, managers and informaticists. In this instance, earlier collaboration with EHR clinical-technical subject matter experts from both acute care and home care would have helped us more realistically determine the feasibility and expense of extracting data across care settings. The importance of engaging these team members early in the research planning cannot be overstated. Sharing a common language with data architects and other technical experts trained in the specifics of the EHR vendor products, the clinical-technical subject matter experts can mitigate many of the challenges we encountered ^31^.

#### 3.2.2 Recommendations

Advancing the collection, extraction and use of data to track physical function to drive clinical decisions, meaningful quality improvement, and clinical research requires significant and thoughtful advanced planning. Recommendations from our experience are highlighted in Table 3.

**Table 3.** Key Recommendations and Considerations for Planning EHR-Based Physical Function Data Collection.

| Recommendation | Examples | Considerations |
| --- | --- | --- |
| 1. Create clear operational definitions for key constructs | Different data elements may describe similar constructs (e.g., pain, physical function) | - Who collects the data?<br>- Frequency of data collection<br>- What tools are used?<br>- What is the quality of the data? |
| 2. Identify specific cohort and associated data elements | All patients vs. patients receiving rehabilitation only | - Lack of clear indicators of admission/discharge/transition<br>- What data elements are expected? |
| 3. Collaborate with those knowledgeable in documentation, EHR access, and billing | Informaticists, managers, clinicians | - Initiate early and maintain ongoing collaboration<br>- Budget for collaborators' time |
| 4. Assess data availability before extraction | Pain, medical status changes, medication use, diagnoses, clinical events (falls, DVTs, pressure ulcers, pneumonia) | - How, when, and by whom is data generated?<br>- Where is it stored in the EHR? |
| 5. Establish a realistic budget and leverage technology | Text-based and other complex data | - Consider natural language processing and machine learning tools |

## 4 CONCLUSIONS

Physical function is a critical and measurable outcome for both clinicians and researchers seeking to improve the value of health care. As a diagnosis-agnostic indicator, physical function offers valuable insights regardless of whether patients receive services from a rehabilitation provider. Declines in physical function can signal increased risk for poor health outcomes, and early recognition creates opportunities to intervene and potentially alter the trajectory of a patient’s health. Standardizing how physical function is measured has become increasingly important—particularly as researchers aim to curate and integrate data across health systems using common data models. In this paper, we highlighted specific challenges and proposed potential solutions for using EHR data to track physical function across care transitions, specifically from acute care to home health care, within a large health system. While clinical data in EHRs are complex and often imperfect, we offer several practical recommendations for future efforts in this space.

## Data Availability

All data produced in the present work are contained in the manuscript.

## Acknowledgements

This project was supported by a pilot grant from the Center on Health Services Training and Research (CoHSTAR). We would also like to express our appreciation of the Core for Clinical Research Data Acquisition (CCDA), a Johns Hopkins Medicine Data Trust analytic team, for the data extraction.

## References

1. Young DL, Fritz JM, Kean J, Thackeray A, Johnson JK, Dummer D, Passek S, Stilphen M, Beck D, Havrilla S, Hoyer EH, Friedman M, Daley K, Marcus RL. Key Data Elements for Longitudinal Tracking of Physical Function: A Modified Delphi Consensus Study. Phys Ther. Apr 1 2022;102(4)doi:10.1093/ptj/pzab279

2. Cummings SR, Studenski S, Ferrucci L. A diagnosis of dismobility--giving mobility clinical visibility: a Mobility Working Group recommendation. JAMA. May 2014;311(20):2061–2. doi:10.1001/jama.2014.3033

3. Musich S, Wang SS, Ruiz J, Hawkins K, Wicker E. The impact of mobility limitations on health outcomes among older adults. Geriatr Nurs. Mar-Apr 2018;39(2):162–169. doi:10.1016/j.gerinurse.2017.08.002

4. Freiberger E, Sieber CC, Kob R. Mobility in Older Community-Dwelling Persons: A Narrative Review. Front Physiol. 2020;11:881. doi:10.3389/fphys.2020.00881

5. Kansagara D, Englander H, Salanitro A, Kagen D, Theobald C, Freeman M, Kripalani S. Risk prediction models for hospital readmission: a systematic review. JAMA. Oct 19 2011;306(15):1688–98. doi:10.1001/jama.2011.1515

6. Krumholz HM. Post-hospital syndrome--an acquired, transient condition of generalized risk. N Engl J Med. Jan 10 2013;368(2):100–2. doi:10.1056/NEJMp1212324

7. Greysen SR, Stijacic Cenzer I, Auerbach AD, Covinsky KE. Functional impairment and hospital readmission in Medicare seniors. JAMA Intern Med. Apr 2015;175(4):559–65. doi:10.1001/jamainternmed.2014.7756

8. Hoyer EH, Needham DM, Atanelov L, Knox B, Friedman M, Brotman DJ. Association of impaired functional status at hospital discharge and subsequent rehospitalization. J Hosp Med. May 2014;9(5):277–82. doi:10.1002/jhm.2152

9. Hardy SE, Kang Y, Studenski SA, Degenholtz HB. Ability to walk 1/4 mile predicts subsequent disability, mortality, and health care costs. J Gen Intern Med. Feb 2011;26(2):130–5. doi:10.1007/s11606-010-1543-2

10. Prevention CfDCa. Persons with hospital stays in the past year, by selected characteristics: United States, selected years 1997–2018. Accessed April 24, 2025, 2025. https://www.cdc.gov/nchs/data/hus/2019/040-508.pdf

11. Boyd CM, Landefeld CS, Counsell SR, Palmer RM, Fortinsky RH, Kresevic D, Burant C, Covinsky KE. Recovery of activities of daily living in older adults after hospitalization for acute medical illness. J Am Geriatr Soc. Dec 2008;56(12):2171–9. doi:10.1111/j.1532-5415.2008.02023.x

12. Leventhal H, Weinman J, Leventhal EA, Phillips LA. Health Psychology: the Search for Pathways between Behavior and Health. Annu Rev Psychol. 2008;59:477–505. doi:10.1146/annurev.psych.59.103006.093643

13. Porter ME, Lee TH. From Volume to Value in Health Care: The Work Begins. JAMA. Sep 13 2016;316(10):1047–8. doi:10.1001/jama.2016.11698

14. Baumhauer JF. Patient-Reported Outcomes - Are They Living Up to Their Potential? N Engl J Med. Jul 6 2017;377(1):6–9. doi:10.1056/NEJMp1702978

15. Ackerly DC, Grabowski DC. Post-acute care reform--beyond the ACA. N Engl J Med. Feb 20 2014;370(8):689–91. doi:10.1056/NEJMp1315350

16. Thomas EM, Smith J, Curry A, Salsberry M, Ridgeway K, Hunt B, Desanto K, Falvey JR. Association of physical function with hospital readmissions among older adults: A systematic review. J Hosp Med. Mar 2025;20(3):277–287. doi:10.1002/jhm.13538

17. Jewell DV, Moore JD, Goldstein MS. Delivering the physical therapy value proposition: a call to action. Phys Ther. Jan 2013;93(1):104–14. doi:10.2522/ptj.20120175

18. Baumhauer JF, Bozic KJ. Value-based Healthcare: Patient-reported Outcomes in Clinical Decision Making. Clin Orthop Relat Res. Jun 2016;474(6):1375–8. doi:10.1007/s11999-016-4813-4

19. Teisberg E, Wallace S, O’Hara S. Defining and Implementing Value-Based Health Care: A Strategic Framework. Acad Med. May 2020;95(5):682–685. doi:10.1097/ACM.0000000000003122

20. Porter ME. What is value in health care? N Engl J Med. Dec 23 2010;363(26):2477–81. doi:10.1056/NEJMp1011024

21. Johnson KB, Neuss MJ, Detmer DE. Electronic health records and clinician burnout: A story of three eras. J Am Med Inform Assoc. Apr 23 2021;28(5):967–973. doi:10.1093/jamia/ocaa274

22. Hoyer EH, Friedman M, Lavezza A, Flanagan E, Kumble S, D’Alessandro M, Gutierrez M, Colantuoni E, Brotman DJ, Young DL. A unit-based, multi-center evaluation of adopting mobility measures and daily mobility goals in the hospital setting. Appl Nurs Res. Apr 2023;70:151655. doi:10.1016/j.apnr.2022.151655

23. Hoyer EH, Friedman M, Lavezza A, Wagner-Kosmakos K, Lewis-Cherry R, Skolnik JL, Byers SP, Atanelov L, Colantuoni E, Brotman DJ, Needham DM. Promoting mobility and reducing length of stay in hospitalized general medicine patients: A quality-improvement project. J Hosp Med. May 2016;11(5):341–7. doi:10.1002/jhm.2546

24. Hoyer EH, Young DL, Klein LM, Kreif J, Shumock K, Hiser S, Friedman M, Lavezza A, Jette A, Chan KS, Needham DM. Toward a Common Language for Measuring Patient Mobility in the Hospital: Reliability and Construct Validity of Interprofessional Mobility Measures. Phys Ther. Feb 1 2018;98(2):133–142. doi:10.1093/ptj/pzx110

25. Klein LM, Young D, Feng D, Lavezza A, Hiser S, Daley KN, Hoyer EH. Increasing patient mobility through an individualized goal-centered hospital mobility program: A quasi-experimental quality improvement project. Nurs Outlook. May-Jun 2018;66(3):254–262. doi:10.1016/j.outlook.2018.02.006

26. McLaughlin KH, Friedman M, Hoyer EH, Kudchadkar S, Flanagan E, Klein L, Daley K, Lavezza A, Schechter N, Young D, Group J-A. The Johns Hopkins Activity and Mobility Promotion Program: A Framework to Increase Activity and Mobility Among Hospitalized Patients. J Nurs Care Qual. Apr-Jun 01 2023;38(2):164–170. doi:10.1097/NCQ.0000000000000678

27. McLaughlin KH, Young D, Friedman LA, Peters J, Vickery G, Hoyer EH. An interprofessional examination of the Johns Hopkins Mobility Goal Calculator among hospitalized postsurgical patients. Nurs Health Sci. Sep 2022;24(3):735–741. doi:10.1111/nhs.12972

28. Young DL, Hannum SM, Engels R, Colantuoni E, Friedman LA, Hoyer EH. Dynamic Prediction of Post-Acute Care Needs for Hospitalized Medicine Patients. J Am Med Dir Assoc. Jul 2024;25(7):104939. doi:10.1016/j.jamda.2024.01.008

29. Qualls LG, Phillips TA, Hammill BG, Topping J, Louzao DM, Brown JS, Curtis LH, Marsolo K. Evaluating Foundational Data Quality in the National Patient-Centered Clinical Research Network (PCORnet(R)). EGEMS (Wash DC). Apr 13 2018;6(1):3. doi:10.5334/egems.199

30. French MA, Hartman P, Hayes HA, Ling L, Magel J, Thackeray A. Coverage of Physical Therapy Assessments in the Observational Medical Outcomes Partnership Common Data Model. Appl Clin Inform. Oct 2024;15(5):1003–1012. doi:10.1055/a-2401-3688

31. Daley KN. Adding Power to Systems Science in Rehabilitation. Phys Ther. Aug 1 2018;98(8):725–726. doi:10.1093/ptj/pzy061

